# Validation of two efficient and robust smartphone-based threshold (GRaBr) and loudness (rACALOS) measures in typical home settings

**DOI:** 10.1101/2024.11.19.24317529

**Authors:** Chen Xu, Lena Schell-Majoor, Birger Kollmeier

## Abstract

Reliable hearing assessment at home can improve accessibility and reduce dependence on in-clinic testing. To be viable, home-based procedures must provide accurate results within short measurement times and remain robust to factors such as ambient noise and variable user attention. This study validated two such procedures—a Graded Response Bracketing method for pure-tone threshold estimation and a reinforced adaptive categorical loudness scaling method for loudness-growth assessment—using remote, smartphone-based testing. Fifteen young adults with normal hearing completed the tasks at home and in the laboratory. Ambient noise levels in home environments were also recorded. Test–retest reliability was assessed by repeating the home measurements on a separate occasion.

Remote measurements closely matched laboratory results, with mean differences below 1 dB for threshold estimation and below 5 dB for loudness scaling. Test–retest differences obtained at home were small, remaining below 2 dB for threshold estimation and below 1 dB for loudness scaling. These findings demonstrate that smartphone-based pure-tone audiometry and loudness-scaling assessments can achieve high accuracy, efficiency, and reliability when using these procedures, provided that basic acoustic-hygiene conditions (e.g., sufficiently low ambient noise) are maintained.

## INTRODUCTION

Pure-tone audiometry, a threshold-based testing approach, is widely used to detect hearing loss and guide audiogram-based hearing aid fitting. Its application for mobile and remote testing has therefore received much interest (see Almufarrij et al., 2022 for a review). In contrast, supra-threshold methods such as categorical loudness scaling (CLS, Hellbrück, 1987; Kollmeier, 1997) to characterize supra-threshold deficits and support loudness-based fitting have received much less attention in the literature on mobile or remote applications (Kopun et al., 2022; Xu et al., 2024a). However, this method appears to be particularly suitable for such applications because it is inherently robust to device calibration offsets (Almufarrij et al., 2022; Xu et al., 2025) due to its parameter estimation relying on level differences rather than on absolute levels. Hence, both of these methods are attractive for mobile and remote testing if they are reliable, robust against disturbances, and provide valid estimates of what would be measured in a clinical setting. The current paper aims at validating refined versions of them that fulfill these requirements.

Accurate estimation of hearing thresholds remains essential for obtaining reliable reference data in both threshold- and supra-threshold-based tests. Previous work examined the effect of experimenter supervision on smartphone-based pure-tone audiometry and adaptive CLS (ACALOS, Brand and Hohmann, 2002) in a sound-attenuated booth with normal-hearing and hearing-impaired listeners, showing no significant supervision effect (Xu et al., 2024b). To further improve threshold estimation with respect to efficiency and robustness against inattention, we developed the graded adaptive response bracketing (GRaBr) method (Xu et al., 2024a), a model-free adaptive procedure that replaces the conventional yes-no paradigm with a two-interval task in which listeners indicate whether they heard none, one, or two sounds. Compared with the single-interval up–down (SIUD) method, which presents a cue tone in a second interval at a fixed level above the target tone (Lecluyse & Meddis, 2009), GRaBr adaptively adjusts this level difference. Simulation studies, including Xu et al. (2024a), showed that this adaptive strategy yields substantially higher efficiency and robustness, particularly under both long- and short-term inattention. However, a validation of the remote GRaBr listening test with real participants in a home environment—where susceptibility to distractions is higher—against laboratory-based assessments is still lacking, which motivated part of the present study.

Furthermore, we consider the adaptive CLS procedure for remote testing which poses challenges in fluctuating ambient noise in home environments that could affect loudness judgments at low sound pressure levels (SPL). Oetting et al. (2014) reported that the mean intra-subject standard deviation of loudness levels close to the threshold was notably high (around 10 dB), yielding significant variability in the hearing threshold estimation from loudness judgments near the threshold. In contrast, the mean intra-subject standard deviation at the level corresponding to 50 categorical units on the loudness function (i.e., the estimated uncomfortable loudness level) was below 5 dB, indicating that this measure is more reliable than the threshold estimate. Hence, despite its advantages for assessing supra-threshold hearing problems mentioned above, the conventional ACALOS procedure often fails to accurately predict audiometric thresholds, likely due to differences in stimulus type and psychophysical paradigm (Oetting et al., 2014). To overcome this limitation, we propose the reinforced adaptive categorical loudness scaling (rACALOS) method (Xu, 2025), which incorporates a refined threshold estimation process into ACALOS. This unified approach combines threshold and supra-threshold assessments in a single, time-efficient test, improving estimation accuracy of the hearing threshold and facilitating remote implementation on smartphones.

Taken together, the present study aims at validating the performance and test-retest reliability of these novel smartphone-based methods for remote hearing assessment - GRaBr for threshold estimation and rACALOS for categorical loudness scaling - when conducted by listeners in typical home environments in comparison to the reference tests conducted inside the laboratory using the baseline procedures. This study evaluates whether these procedures yield accurate and reliable measures under realistic, uncontrolled acoustic conditions which is a prerequisite for future integration into mobile hearing assessment platforms.

## METHODS

### Participants

Fifteen young adults with normal hearing (aged 20 to 35 years; 4 men, 11 women) participated in this study. All participants were members of working groups or students at the University of Oldenburg and were recruited primarily through informal requests within the labs. The three authors did not participate in the study as subjects. All participants self-reported no hearing issues and were classified as being normal hearing (NH) in former studies. Two inclusion criteria were applied: (i) the air-conduction pure-tone average (PTA-4) at 0.5, 1, 2, and 4 kHz in the better ear had to be less than or equal to 20 dB HL, and (ii) symmetric hearing, defined as a threshold difference of no more than 20 dB between ears at any test frequency. All 15 participants met these criteria. Some listeners (N = 5) received compensation of €12 per hour for their participation, while others took part as part of their work duties. The study was approved by the Research Ethics Committee of the University of Oldenburg (Drs. EK/2023/004).

### Reference Laboratory-Based Testing Inside a Booth

We reused the data set from our previous study (Xu et al., 2024b) as the reference laboratory-based listening tests. Full methodological details are provided in Xu et al. (2024b). Following a repeated-measures design, the participants in the present study were the same individuals as those in Xu et al. (2024b). All measurements were conducted by an experimenter inside a sound-attenuated booth. Hearing thresholds were obtained using the SIUD procedure (Lecluyse & Meddis, 2009), and loudness growth functions were assessed using the ACALOS procedure (Brand & Hohmann, 2002). Please note that the SIUD procedure has been validated against the clinical audiograms in Lecluyse and Meddis (2009), hence we assume that thresholds measured by SIUD are comparable to the standard clinical audiogram. Both tests were performed at 0.25, 1, and 4 kHz.

### Equipment, Procedure, and Environment for Remote Testing

Prior to the start of remote testing, a test kit was assembled, which included a smartphone (OnePlus, Android), a USB-C charger, and HD650 circumaural headphones (Sennheiser, Wedemark, Hanover, Germany). The smartphone and headphones were pre-calibrated using a Brüel & Kjær (B&K) artificial ear 4153, a B&K 0.5-inch microphone 4134, a B&K microphone pre-amplifier 2669, and a B&K measuring amplifier 2610, with a target calibration level of 80 dB SPL. There was only one test kit in use, and the setup was calibrated once at the beginning of data collection. Upon handing over the test kit, participants received a brief oral explanation of the remote experiments, and consent forms were signed before they began. The oral instructions consisted of three parts: test environment check, internet connection check, and steps for conducting the remote experiments. The details are provided below. Participants could initiate testing at home by connecting to the internet via WLAN and accessing the provided website. For data security, a VPN connection was established using the ‘GlobalProtect’ app when accessing the site. The workflow of the web-based application for remote testing was described in Xu et al. (2024b). A Raspberry Pi 3 Model B (Raspberry Pi Foundation, UK), a Linux-based micro-controller, served as the server hosting the measurement site. All behavioral data were stored on an SD card within the Raspberry Pi, located at the University of Oldenburg.

Listeners were asked to complete remote testing at home on two separate days within a week—one for the test measurement and the other for the retest measurement. Each day’s testing lasted approximately half an hour, resulting in a total testing time of one hour per participant.

The home environments were primarily located in rural regions of northwestern Germany and cities such as Oldenburg, Cloppenburg, Jever, and Bad Zwischenahn.

### Noise Level Measurement for Remote Testing

Ambient noise was measured using the freely available “Decibel X” smartphone app (SkyPaw Co., Ltd.), configured with an A-weighted filter and a 500-ms slow time weighting. The app displayed real-time, average, and maximum noise levels but did not record audio. To calibrate the smartphone’s internal microphone, a Class 3 digital sound level meter (Voltcraft SL-100; ±2 dB at 1 kHz), previously calibrated in our laboratory, was placed next to the phone while 80-dB SPL narrowband noise was presented via a laptop. The app’s gain was then adjusted to match the digital meter, requiring a correction of 13.7 dB. The same smartphone and headphones were used for all participants to ensure consistent gain.

At the beginning of each remote session, participants documented the current ambient noise level. They were instructed to monitor real-time noise using the app and to pause testing whenever levels exceeded 45 dB(A), a threshold selected based on Kopun et al. (2022), who showed that remote loudness-scaling results remain comparable to laboratory measurements below 50 dB(A) ambient noise level. The time and location of each remote session were also recorded.

### Remote Listening Tests at Home

A total of 24 sessions were conducted by each subject at home, consisting of 4 listening tests (SIUD, GRaBr, ACALOS, and rACALOS; see details below) across 3 test frequencies (0.25, 1, and 4 kHz) and 2 runs (test and retest), presented in randomized order. Please note that the test and retest measurements are referred to as Run 1 and Run 2, respectively. Participants were instructed to take short breaks between sessions.

#### Remote pure-tone audiometry

Remote pure-tone thresholds were measured using two adaptive procedures: the SIUD method and the GRaBr approach (Lecluyse et al., 2009; Xu et al., 2024a). In both tasks, each trial contained a probe tone and a higher-level cue tone, and listeners reported whether they heard zero, one, or two tones. The cue tone was muted on 20% of trials (catch trials). The key difference between the procedures is that SIUD uses a fixed 10-dB level difference between probe and cue tones, whereas GRaBr adapts this difference: it starts at 10 dB, decreases to 5 dB after the first “one-tone” response, and to 2 dB after the second.

In SIUD, the step size of the probe tone level began at 10 dB and was reduced to 2 dB after the first “one-tone” response (by setting the level to the midpoint of the preceding two levels). In GRaBr, step sizes were 8, 6, 3, and finally 1 dB after the first, second, and third reversals, respectively. In both procedures, levels were adjusted adaptively—becoming harder after correct responses and easier after incorrect ones. To enable fair comparison, key parameters (e.g., minimum trials, reversals, starting level) were matched. The initial cue-tone level for both methods was drawn from a discrete uniform distribution of 55–65 dB SPL (1-dB steps). Each track ended after at least 14 reversals and 10 trials, and the first four reversals were discarded.

Each pure tone lasted 0.2 s, with cosine ramps of 0.02 s and a 0.3 s interval between tones. In SIUD, the correct response rates were fitted to an S-shaped logistic psychometric function, and the level at the 50% correct response point (L_50_) was estimated as the hearing threshold. For GRaBr, responses from the upper and lower tracks (i.e., the tracks of the cue tones and the probe tones) were fitted to two independent psychometric functions, and the hearing threshold was calculated as the mean level at the 50% correct response point of both functions (i.e., 0.5*(L_50,upper_ + L_50,lower_)).

#### Remote adaptive categorical loudness scaling

The adaptive categorical loudness scaling (ACALOS) method was used to assess the loudness growth function (Brand & Hohmann, 2002; ISO 16832, 2006). In the ACALOS task, participants rated the loudness of stimuli on an 11-point scale with descriptors ranging from ‘very soft”, “soft”, “medium”, “loud”, and “very loud” with 4 unnamed intermediate categories in between, plus the two limiting categories “not heard”, and “too loud”. The stimulus levels, ranging from -10 to 105 dB, were presented in a pseudo-random order following an initial estimation of the user-specific dynamic range (Phase I, see Fig. 1), which was updated to obtain a more representative placement of test level in Phase II, encompassing 26 trials. At the end of the procedure, a loudness growth function was modeled by fitting two linear segments and a transition region using a Bezier fit, following the BTUX fitting method (Oetting et al., 2014). Each frequency was assessed in a separate run.

**Fig. 1.**
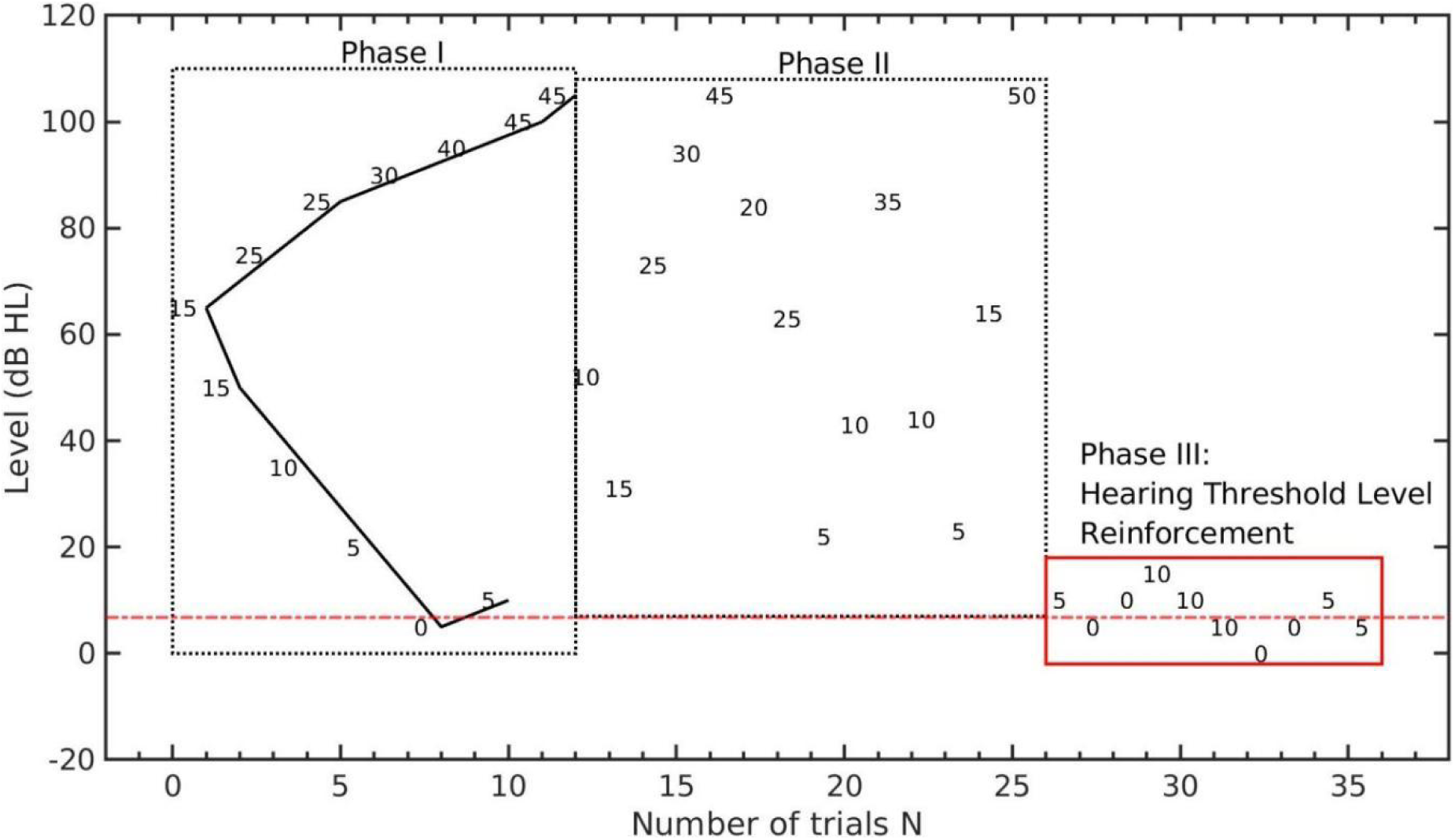
An example track of the reinforced adaptive categorical loudness scaling (rACALOS), where the level (in dB HL) is plotted as a function of the number of trials N. The listener’s response (in categorical units (CU)) is annotated with numbers between 0 (‘not heard’) and 50 (‘too loud’). Left dotted rectangle region: Phase I (‘dynamic range estimation’); Middle dotted rectangle region: Phase II (‘presenting and re-estimation’); Right solid red rectangle region: Phase III (‘hearing threshold level reinforcement’); Red dash-dotted line: target threshold. In Phase III, the step size is set to 5 dB, and the number of trials is set to 10.

An example run of the proposed rACALOS procedure is shown in Fig. 1. The rACALOS followed the same adaptive rules as ACALOS during Phases I and II (see above) but presented additional stimuli near the hearing threshold to better estimate HTL. The starting level of Phase III was set at the minimum level reached in Phases I and II, plus 5 dB. In this phase, a one-up-one-down adaptive rule was applied: the stimulus level increased by 5 dB if participants responded with “not heard” and decreased by 5 dB if they selected other loudness categories (e.g., “very soft,” “medium”). Phase III consisted of 10 trials. The stimuli used were one-third-octave-band low-noise noises (Kohlrausch et al., 1997). Each noise stimulus had a duration of 1 second with 0.05-second rise and fall ramps.

## RESULTS

### Noise Level Measurements

Fig. 2 presents a box plot of the ambient noise levels recorded by each participant (N = 15), who documented the noise level a total of 24 times at the start of each measurement session, corresponding to 24 measurement sessions at home within a week. Notably, the noise levels for all participants remained below the recommended upper limit of 45 dB(A). The median noise level across subjects was 36.0 dB, which was approximately 0.5 dB higher than the reference noise level measured inside the sound-attenuated booth. The interquartile range (IQR) across participants was 1.2 dB. Overall, the sound levels in participants’ homes were considerably low and comparable to those measured within the booth, indicating a suitable test environment. A few participants (e.g., No. 2 and No. 8) lived near a train station, resulting in slightly elevated noise levels compared to others. Additionally, one participant (No. 1) misinterpreted the task and consistently rounded the recorded noise level to an integer, leading to uniform values across sessions.

**Fig. 2.**
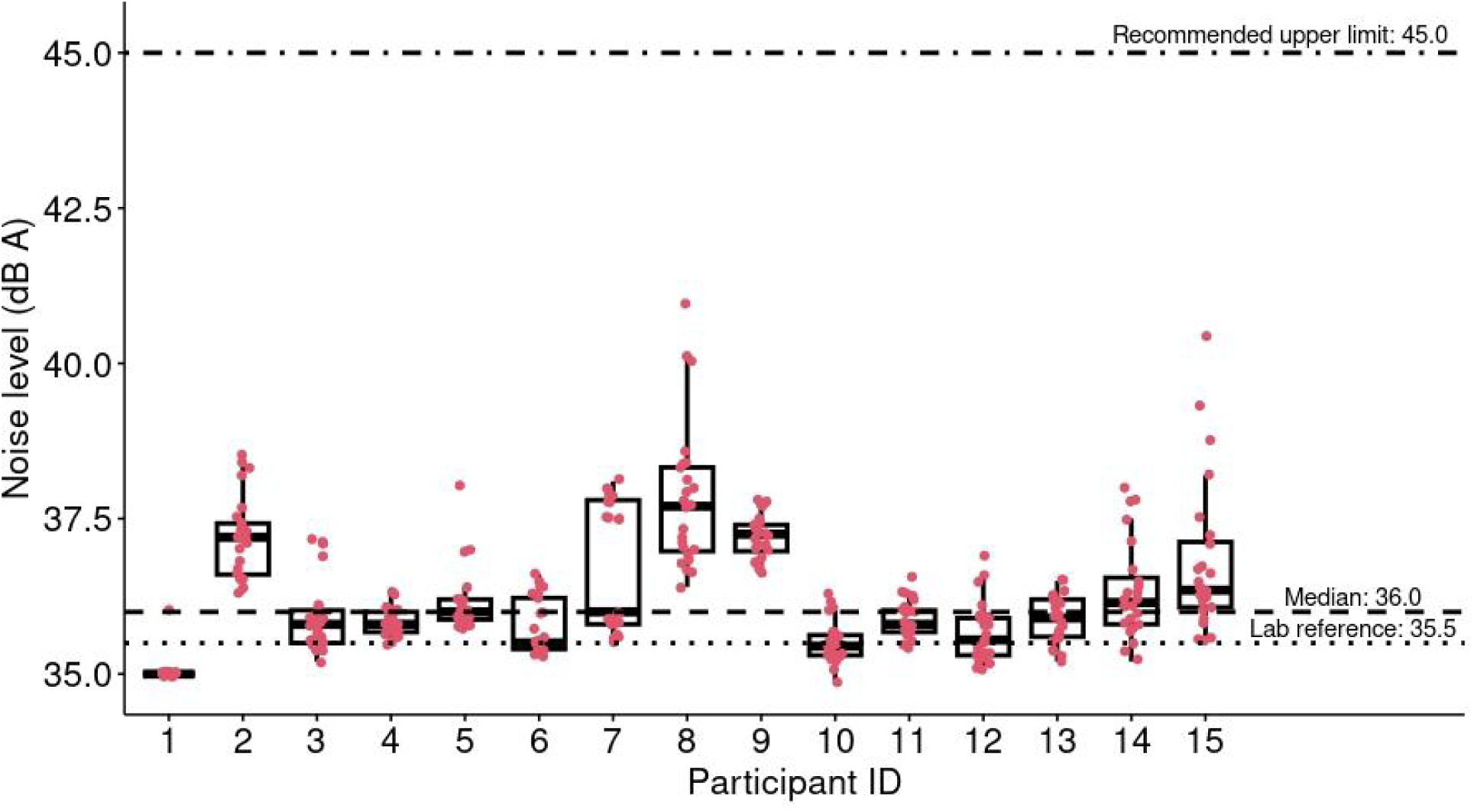
Ambient noise level (in dB A) measurement across participants (N = 15). Medians, 25th and 75th percentiles, and interquartile ranges (IQR) are visualized in the box-plot while the end of the whiskers denotes the minimum and maximum, indicating the 5th and 95th percentiles respectively. Circles represent individual data points. Dotted line: lab reference (i.e., ambient noise level measured within a booth). Dashed line: median value across subjects. Dot-dashed line: recommended upper limit.

### Validation of the GRaBr procedure in a home environment

To validate the home-based GRaBr procedure, its thresholds were compared with booth-based thresholds obtained using the SIUD method at 0.25, 1, and 4 kHz. As reported in Table 1, threshold differences were normally distributed (Shapiro–Wilk p = 0.92) and showed a negligible mean bias of 0.4 dB. The correlation between home and booth measurements was moderate—likely reflecting the limited variability in this normal-hearing sample—and the RMSE was 7 dB, consistent with previous smartphone-based audiometry studies. Therefore, the GRaBr procedure for home testing is comparable to the reference in-lab threshold measurements, suggesting good validity.

**Table 1.** Validation of the home-based GRaBr procedure for threshold measurements against the in-lab reference measurement using SIUD. Differences in hearing thresholds between the two measurements are shown. Spearman correlation coefficients (R), root-mean-square error (RMSE), and bias are reported for each of the three test frequencies and for their overall average.

| Frequency (Hz) | 250 | 1000 | 4000 | overall |
| --- | --- | --- | --- | --- |
| R (“spearman”) | 0.29 | 0.61 | 0.29 | 0.47 |
| RMSE (dB) | 8.0 | 4.6 | 7.9 | 7.0 |

| Bias (dB) | 5.5 | 1.0 | -5.3 | 0.4 |
| --- | --- | --- | --- | --- |

A two-way repeated-measures ANOVA showed no main effect of test environment (F(1,14) = 0.09, p = 0.77). Frequency had a significant effect (F(2,28) = 20.80, p < 0.05), as did the environment × frequency interaction (F(2,28) = 36.01, p < 0.05). Post-hoc tests revealed no significant difference between home and booth thresholds at 1 kHz, while small but significant differences were found at 0.25 and 4 kHz.

### Validation of the rACALOS procedure in a home environment

Loudness growth functions measured at home using rACALOS were compared with those obtained in the booth using a standard ACALOS procedure at 0.25, 1, and 4 kHz. Differences followed a normal distribution (Shapiro–Wilk p = 0.22), and the overall bias across categorical units (CUs) was small (3.38 dB), indicating good validity of rACALOS relative to standard ACALOS. Agreement metrics varied across loudness levels: correlations for CUs ≥ 35 ranged from 0.57 to 0.62, whereas correlations for softer CUs were lower (< 0.45), accompanied by larger RMSE values. This pattern reflects the higher variability inherent at near-threshold levels rather than a systematic effect of home-environment noise, as deviations occurred in both directions. See Table S1 in the supplementary material for the details of the validation for the home-based rACALOS procedure.

A three-way repeated-measures ANOVA assessing test environment, frequency, and CU revealed no significant main effect of environment. Frequency (F(2,28) = 5.33, p < 0.05) and CU (F(3,38) = 353.30, p < 0.05) showed significant effects. Post-hoc comparisons indicated that environment-related differences emerged only at a few specific combinations (4 kHz at 5, 25, and 45 CU), while the majority of frequency–CU combinations showed no significant differences.

### Test-Retest Reliability for home testing

To assess the reliability of the four home-based listening tests, measurements from the initial run (run 1) were compared with those from the retest (run 2). Details of the statistical analyses in terms of reliability for remote ACALOS and rACALOS testing are provided in the supplementary material (see Table S2).

#### SIUD and GRaBr for home testing

As shown in Table 2, the GRaBr procedure showed test-retest intraclass correlation coefficient (ICC) values exceeding 0.75 (p < 0.05), indicating good reliability across all three frequencies, whereas the SIUD procedure yielded ICC values ranging from 0.59 to 0.77 (p < 0.05), reflecting moderate test-retest reliability. This difference was significant (p < 0.05), i.e., GRaBr demonstrated significantly higher test-retest reliability than SIUD based on these metrics.

**Table 2.** Test-retest reliably between two runs for remote GRaBr and SIUD testing at 0.25, 1, and 4 kHz frequencies. Correlation coefficient (both ‘Pearson’ and ‘Spearman’), RMSE, bias, and intraclass cross-correlation (ICC) between the test and retest measurements are reported.

|  | GRaBr (N = 15) |  |  | SIUD (N = 15) |  |  |
| --- | --- | --- | --- | --- | --- | --- |
| Frequency (Hz) | 250 | 1000 | 4000 | 250 | 1000 | 4000 |
| R (Pearson) | 0.70 | 0.90 | 0.73 | 0.43 | 0.61 | 0.63 |
| R (Spearman) | 0.47 | 0.78 | 0.65 | 0.40 | 0.57 | 0.65 |
| RMSE | 4.0 | 3.3 | 3.4 | 5.4 | 5.2 | 4.1 |
| Bias | -1.5 | 2.0 | -1.3 | -1.7 | -0.5 | -1.4 |
| ICC2k<sup>a</sup> | 0.77 | 0.88 | 0.83 | 0.59 | 0.77 | 0.74 |

<sup>a</sup> ICC2k: Average random raters
<sup>b</sup> ns: not significant; \* $p < 0.05$ ; \*\* $p < 0.01$ ; \*\*\* $p < 0.001$ ; \*\*\*\* $p < 0.0001$

A significant main effect of frequency was observed (F(2, 28) = 15.46, p < 0.05). Moreover, pairwise t-tests were performed to assess reliability by comparing the two runs for both adaptive procedures across all three frequencies, showing no significant differences (bias) between runs in most cases, except for GRaBr at 1 kHz (t(14) = 2.85, p < 0.05).

#### ACALOS and rACALOS for home testing

The reliability of the ACALOS and rACALOS procedures was assessed using across-run bias (quantified by mean signed difference, MSD) and within-run variability (measured by mean interquartile range, MIQR) (please see Kopun et al. (2022) for the definitions of the MSD and MIQR). Both adaptive procedures demonstrated an MSD of less than 5 dB at all frequencies, indicating a small across-run bias. Most MIQR values did not exceed 10 dB for either procedure at the three frequencies, although they were typically larger than 10 dB at 5, 10, and 15 CU, reflecting a consistent within-run variability. Overall, these metrics suggested that both ACALOS and rACALOS exhibited strong reliability.

A repeated measures ANOVA revealed a significant main effect of the procedure, indicating a statistically significant difference between ACALOS and rACALOS (p < 0.05). Compared to the ACALOS procedure, the rACALOS procedure yielded lower mean sound levels (by 1.9 dB) across participants and CU. Since the rACALOS and ACALOS procedures are identical in Phases I and II, this difference is likely attributable to the additional trials included in Phase III of the rACALOS procedure (see Fig. 1).

No significant effect was found for frequency (F(2, 28) = 2.51, p = 0.10), and as expected, the two runs (test and retest measurements) did not differ (F(1, 14) = 1.97, p = 0.18). A subsequent post-hoc t-test compared median levels of the ACALOS and rACALOS procedures between runs 1 and 2 across three frequencies and 11 categories, indicating that median levels for run 1 did not significantly differ from those for run 2 in most cases (31 out of 33 groups of comparison = 3 levels of frequency * 11 levels of CU), except for two groups (measurements at 0.25 kHz for 25CU (t(29) = 2.08) and 40 CU (t(29) = 2.21)).

### Accuracy of HTL Estimation for the rACALOS procedure

Pure-tone audiometric thresholds are plotted against CLS thresholds (i.e., levels at 2.5 CU of the loudness growth functions) for two runs and three frequencies in Fig. 3. Compared to ACALOS, the majority of rACALOS points were consistently and closely clustered around the diagonal line, indicating that thresholds estimated by the rACALOS method aligned more closely with pure-tone thresholds than those from baseline ACALOS and, hence, provide improved accuracy in threshold estimation. Quantitatively, R values increased by 36% for GRaBr and 23% for SIUD when ACALOS was reinforced near the hearing threshold level.

**Fig. 3.**
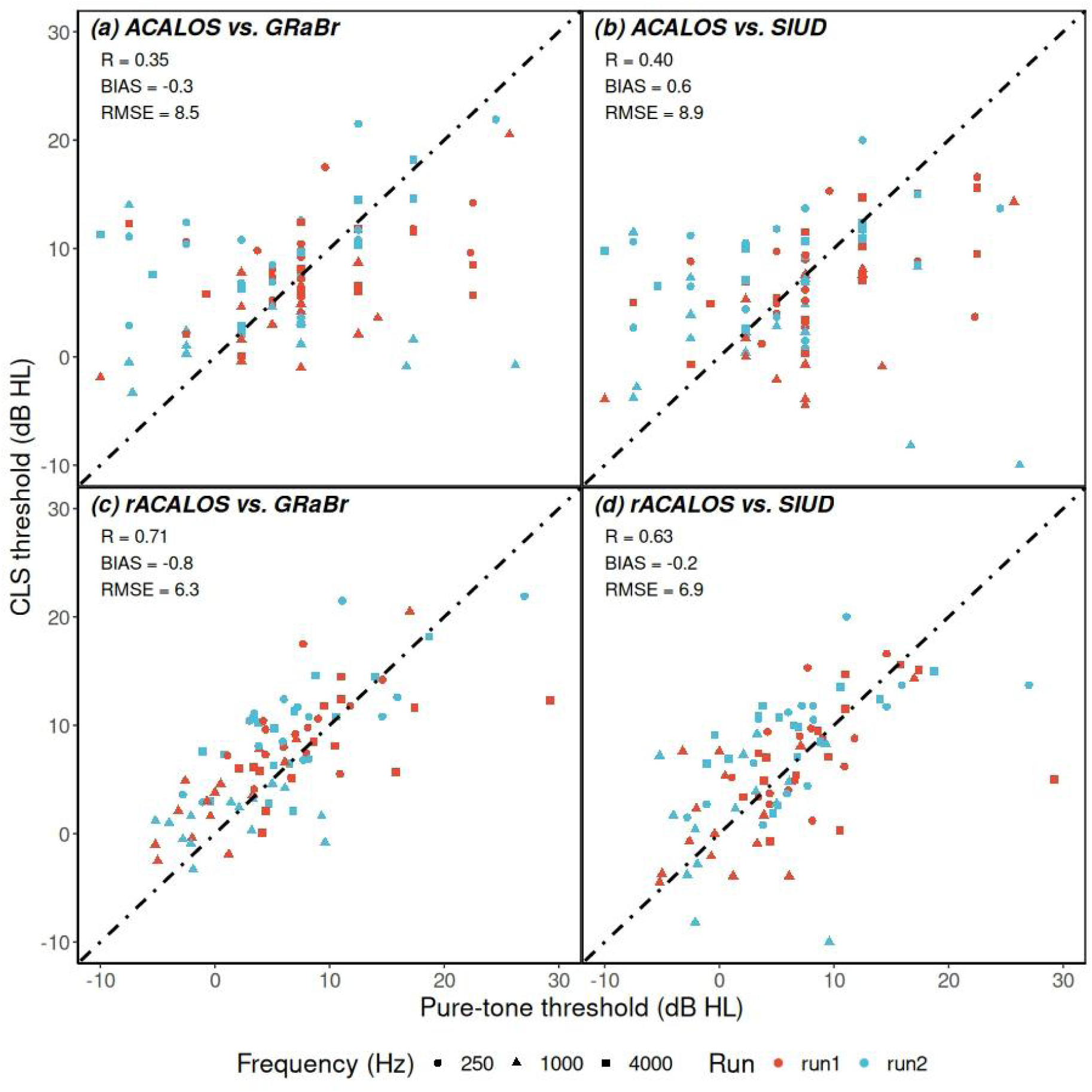
Scatter plot for comparison between pure-tone (abscissa) and CLS (ordinate) thresholds (defined as the level at 2.5 CU of the loudness growth functions) in dB HL of N = 15 individual listeners. Frequency is labeled with different shapes while the run is denoted with different colors (run1: red, run2: blue). A set of statistical metrics (R, Bias, and RMSE) are reported in the top-left corner. For rACALOS, 10 additional trials with a step size of 5 dB were used.

Additionally, RMSE values for the rACALOS method decreased by approximately 2 dB compared to the baseline, while biases remained unchanged. Overall, the reinforcement of baseline ACALOS positively influenced cross-correlation and reduced error.

The highest correlation coefficient and lowest RMSE were observed between GRaBr thresholds and rACALOS, followed by SIUD and rACALOS. In contrast, the unmodified ACALOS procedure showed lower correlation coefficients and higher RMSEs for both threshold estimation methods, indicating the superior performance of rACALOS, as confirmed by t-tests (p < 0.05).

## DISCUSSION

This study validated the performance of two novel smartphone-based procedures—GRaBr for pure-tone audiometry and rACALOS for categorical loudness scaling—conducted by listeners independently in typical home environments. The results demonstrate that accurate and reliable measurements can be obtained without professional supervision or sound-treated booths, confirming the feasibility of remote hearing assessments using standard smartphones.

### Ambient Sound Levels in Home Settings

The median background level measured across participants’ homes was 36.0 dB(A), comparable to quiet residential environments and well below limits defined by ANSI S3.1–1999 (R2018) for covered-ear conditions. These values fall within the updated permissible ranges for circumaural earphones proposed by Margolis et al. (2022), ensuring that ambient sound did not measurably affect the results. While noise levels were not experimentally manipulated, participants were advised to continuously monitor them to confirm that real-world domestic conditions remained suitable for valid measurements. Consequently, the obtained home results were closely aligned with those from the booth.

The recorded noise levels were lower than those in earlier remote audiometry studies (Storey et al., 2014; Brennan-Jones et al., 2016; Swanepoel et al., 2015) and comparable to the quietest non-sound-treated settings reported by Serpanos et al. (2022) and Bean et al. (2022). It is likely that our participants conducted the smartphone-based listening tests at home in rural areas during the morning or evening, whereas other studies typically test in clinical settings located in urban areas during the daytime, which tend to be noisier. Overall, the measured noise conditions confirm that typical homes can provide sufficiently quiet environments for reliable mobile hearing assessments.

### Pure-Tone Audiometry (GRaBr)

Smartphone-based audiometry with GRaBr showed good agreement with in-booth reference measurements, with a mean bias of 0.4 dB. While both SIUD (0.59 < ICC < 0.77) and GRaBr performed reliably, GRaBr achieved higher test–retest consistency (ICC > 0.75, p < 0.05) and lower within-subject variability, confirming its improved stability predicted by simulation (Xu et al., 2024a). These findings extend prior validations of boothless audiometry (Maclennan-Smith et al., 2013; Swanepoel et al., 2015; Serpanos et al., 2022) to an unsupervised, mobile context.

The correlation between at-home and in-booth thresholds (R = 0.47) was lower than in studies involving hearing-impaired listeners (R > 0.9; Maclennan-Smith et al., 2013), likely because our normal-hearing sample exhibited limited threshold variability. Future refinements - such as automated quality control or remote calibration - may further optimize the GRaBr for remote threshold testing.

### (reinforced) Adaptive Categorical Loudness Scaling

Both ACALOS and its reinforced version, rACALOS, yielded valid and reliable loudness functions in typical home conditions. The bias between in-booth and at-home measurements (3.4 dB) was smaller than that reported by Kopun et al. (2022), indicating strong agreement across environments. The enhanced rACALOS design produced smaller within-run (IQR) and across-run (MSD) variability than both baseline ACALOS and previously published CLS methods (Rasetshwane et al., 2015; Fultz et al., 2020; Kopun et al., 2022). At 5 CU, the mean IQR values for rACALOS (7.7 dB at 1 kHz and 6.7 dB at 4 kHz) were notably lower than those in earlier studies (≈10–15 dB), reflecting improved stability near the hearing threshold.

Across-run bias was also smaller than in prior work, likely due to the lower average ambient noise level and the reinforced threshold estimation sequence. The additional trials near threshold in rACALOS substantially reduced estimation uncertainty, resulting in the smallest overall variability among tested methods. Compared with alternative adaptive CLS frameworks (Fultz et al., 2020), rACALOS offers higher precision and efficiency, validating its use as a robust supra-threshold measurement for mobile and unsupervised hearing assessments.

### Accuracy of HTL Estimation

Table 3 presents a comparison between our current study and several state-of-the-art works (Fultz et al., 2020; Trevino et al., 2016; Sanchez-Lopez et al., 2021) by evaluating the cross-correlation between CLS and pure-tone thresholds. Multiple CLS methods, including Fixed-level (FL), Maximum expected information-Median (MEL-Med), Maximum expected information-Maximum likelihood (MEL-ML), Slope-adaptive (SA), ACALOS, and rACALOS, were used to estimate thresholds, which were then compared with pure-tone thresholds measured using various audiometric methods such as a clinical audiometer, SIUD, and GRaBr. In the studies by Fultz et al. (2020) and Trevino et al. (2016), correlation coefficients R values (according to “spearman”) ranged from 0.21 to 0.26 for the threshold estimated from all four CLS methods, indicating a relatively weak cross-correlation. Additionally, the RMSEs and biases in these studies were notably large, suggesting that CLS thresholds did not align well with pure-tone thresholds. In contrast, Sanchez-Lopez et al. (2021) applied a baseline ACALOS method using the same audiometric procedure as Fultz et al. (2020), and while the R-value did not significantly improve, both RMSE and bias were notably reduced. In our study, we employed SIUD and GRaBr to measure pure-tone thresholds, yielding a stronger cross-correlation and smaller bias, although the RMSE was slightly larger or comparable to that reported by Sanchez-Lopez et al. (2021).

**Table 3.**
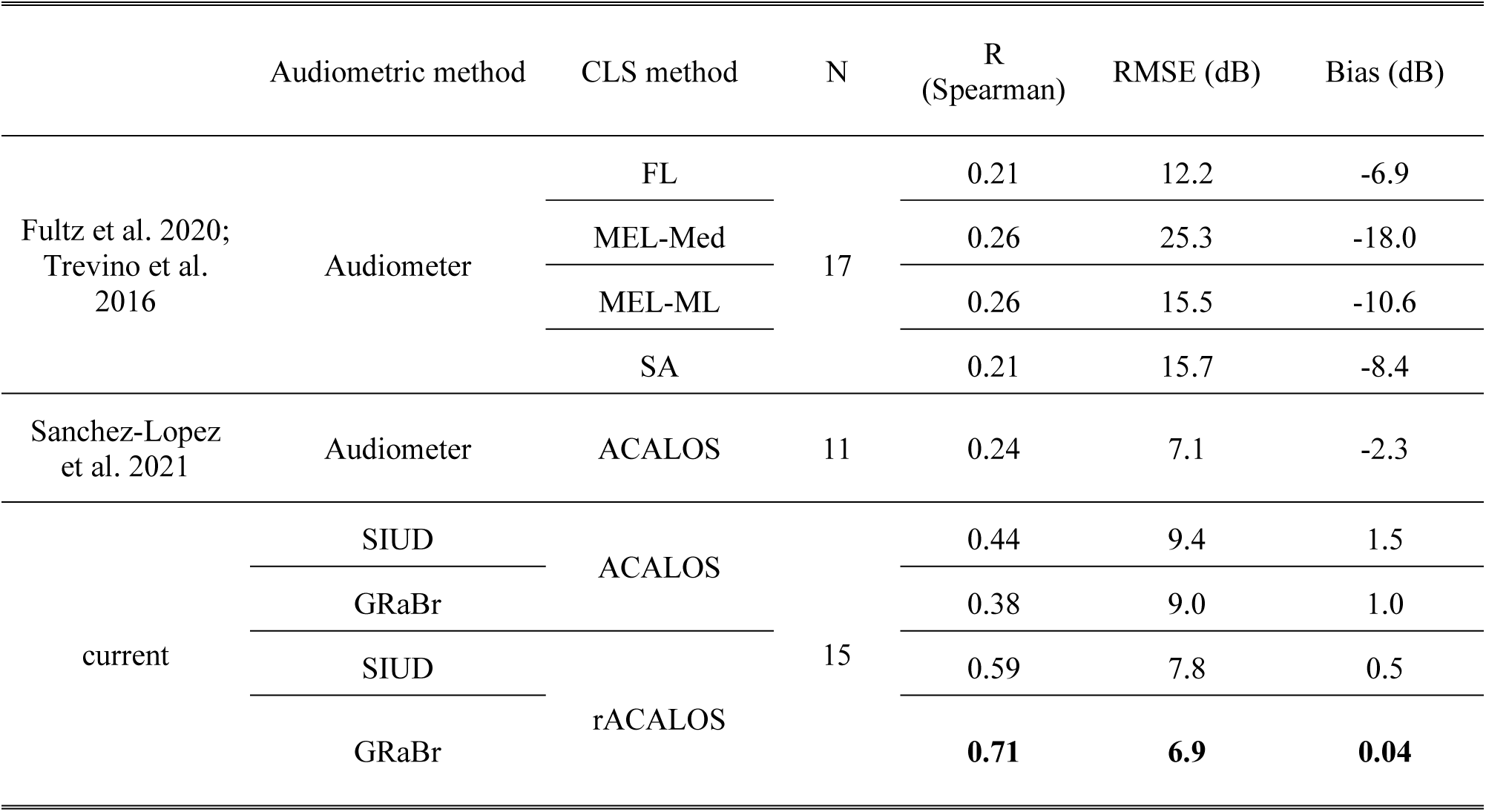
Comparison of threshold estimates obtained from different pure-tone audiometry methods and categorical loudness scaling (CLS) methods across several state-of-the-art studies and the present study. Spearman correlation coefficients (R), root-mean-square error (RMSE), bias, and sample size (N) are reported for each method combination. The highest R value and the lowest RMSE and bias within each comparison set are highlighted in bold. (FL = Fixed-level; MEL-Med = Maximum-expected-information, median; MEL-ML = Maximum-expected-information, maximum likelihood; SA = Slope-adaptive, as defined in Fultz et al. (2020))

|  | Audiometric method | CLS method | N | R<br>(Spearman) | RMSE (dB) | Bias (dB) |
| --- | --- | --- | --- | --- | --- | --- |
| Fultz et al. 2020;<br>Trevino et al.<br>2016 | Audiometer | FL | 17 | 0.21 | 12.2 | -6.9 |
|  |  | MEL-Med |  | 0.26 | 25.3 | -18.0 |
|  |  | MEL-ML |  | 0.26 | 15.5 | -10.6 |
|  |  | SA |  | 0.21 | 15.7 | -8.4 |
| Sanchez-Lopez<br>et al. 2021 | Audiometer | ACALOS | 11 | 0.24 | 7.1 | -2.3 |
| current | SIUD | ACALOS | 15 | 0.44 | 9.4 | 1.5 |
|  | GRaBr |  |  | 0.38 | 9.0 | 1.0 |
|  | SIUD | rACALOS |  | 0.59 | 7.8 | 0.5 |
|  | GRaBr |  |  | **0.71** | **6.9** | **0.04** |

Considering all the studies, the rACALOS method consistently produces thresholds closest to pure-tone thresholds, outperforming other CLS methods. This finding is consistent with computer simulations that examined threshold variability across different parameter combinations of rACALOS compared to the original ACALOS procedure. However, it is important to note that rACALOS requires more measurement time due to the increased number of trials focused on converging near the HTL. Additionally, while pure-tone thresholds obtained using clinical audiometers are still widely regarded as the ‘gold standard’, more precise (see Xu et al., 2024a; Lecluyse & Meddis., 2009) and criterion-free methods such as SIUD and GRaBr may produce stronger correlations with CLS thresholds. It is also crucial to recognize that this comparison is based on a small sample of young NH listeners, and the conclusions may differ if HI listeners are included or if a larger participant pool is studied. This consideration is particularly relevant for potential discrepancies between the narrowband noise thresholds estimated by the CLS methods used here and the pulsed pure-tone thresholds assessed via audiograms. While threshold differences in our study sample of young NH listeners were minimal, variations in stimulus characteristics—such as spectral extent and modulation spectrum—may yield threshold differences in naïve listeners with hearing impairments.

Nonetheless, we also expect these differences to be minimal, as the low-noise, third-octave-band noise utilized here shares key perceptual characteristics with a frequency-modulated sinusoid with minor envelope fluctuations and an instantaneous frequency confined well within a critical band (cf. Zwicker & Fastl, 2013).

### Advantages of rACALOS

#### Increased time efficiency

The rACALOS procedure combines two listening tests—pure-tone audiometry and ACALOS—into a single, integrated protocol. This approach significantly reduces the measurement time required for participants by eliminating the need for separate tests with separate instructions and training procedures.

#### Improved HTL accuracy

Compared to the original ACALOS, rACALOS includes additional trials near the hearing threshold level (HTL), enhancing the precision of HTL estimation (see Table 3 for details). These modifications enable the seamless integration of audiometric measurement into the ACALOS framework.

### Consistent user interface and minimal training requirements

The rACALOS procedure employs the same interface and measurement paradigm as ACALOS. Consequently, participants already familiar with ACALOS require no additional training, while new users need to become familiar with only a single, consistent interface for both listening tests.

### Limitations and Outlook

In this study, we conducted smartphone-based listening tests outside of a sound booth, preceded by ambient noise level measurements. Given the recommendation for conducting tests in rather quiet acoustical conditions, the testing environments generally exhibited a low background noise level. However, many individuals live in urban regions with significant vehicle or industrial noise, where real-world environments are typically much noisier. Testing in such noisy conditions warrants further investigation. Potential solutions, such as circumaural muffs or noise-canceling earphones (NCE), could prove effective. For instance, Saliba et al. (2017) evaluated mobile-based audiometry under 50 dB A background noise, using passive and active noise cancellation by placing circumaural muffs over insert headphones, successfully reducing noise. Similarly, Clark et al. (2017) tested NCE (BoseQuietComfort 15) in a patient consultation room and found that NCE sufficiently attenuated ambient noise below the ANSI standards.

Another key concern for out-of-booth audiometric tests is distraction. As noted by Margolis et al. (2022), background noise not only causes direct masking but also acts as a source of distraction. Their study demonstrated that increasing background noise levels led to elevated hearing thresholds and higher subjective ratings of distraction. Xu et al. (2024a) further supported these findings, characterizing distraction from internal noise (e.g., background noise) as long-term inattention. They also proposed and simulated short-term inattention—where listeners are distracted by external events—during mobile hearing tests, though this has yet to be validated with human participants.

Another limitation of this study is the use of an integrated microphone for noise measurement.

Studies like Kopun et al. (2022) recommend using an external microphone, such as the MicW iBoundary, which provides higher accuracy in capturing frequency characteristics and calibration precision compared to the internal microphone used here.

Enhanced calibration of smartphone microphones can, in principle, be achieved using an external reference sound, such as a whistle tone produced by a standard empty beer bottle (Scharf et al., 2024). However, accurate calibration of the playback level remains a challenge and is essential for precise auditory measurements. While pragmatic approaches such as biological calibration—using a normal-hearing reference listener to define 0 dB HL (Honeth, 2010; Masalski et al., 2014)—may offer a practical alternative, this aspect was beyond the scope of the present proof-of-concept study with its limited sample size. Future work could focus on data-driven estimation of calibration coefficients to enable fully unsupervised field applications (Xu et al., 2025).

In the future, we plan to expand our study by increasing the number of normal-hearing (NH) participants and incorporating hearing-impaired (HI) participants. Compared to NH listeners, we expect the validity of pure-tone audiometry and ACALOS tests in HI listeners to be comparable, as supported by previous studies (e.g., Hazan et al., 2022; Bean et al., 2022; Xu et al., 2024b), suggesting that hearing loss does not significantly affect test validity. Regarding reliability, HI listeners are expected to show similar or sometimes even higher test-retest reliability in audiometry than NH listeners (Hazan et al., 2022), likely due to their elevated hearing thresholds, which reduce the impact of ambient noise. Similarly, in the ACALOS procedure, HI listeners should exhibit similar or even greater reliability at lower levels of the loudness growth function, as they are less affected by background noise. This may have a limited impact on the accuracy of the loudness growth slope fitted to the data, which is generally increased in hearing-impaired listeners exhibiting recruitment. However, since the slope estimate is derived from loudness judgments across multiple supra-threshold levels, it is considered a reliable and robust measure, even when obtained through self-administered smartphone assessments.

Finally, Shen et al. (2018) and Kursun et al. (2023) introduced a quick categorical loudness scaling (qCLS) procedure based on a Bayesian adaptive method, which can estimate equal loudness contours within just 5 minutes. Given its efficiency and accuracy, incorporating qCLS into future smartphone-based loudness tests is worth considering. However, it remains uncertain whether qCLS can estimate hearing thresholds as precisely as the rACALOS developed in this study, highlighting the need for further research to evaluate its threshold accuracy in comparison.

## CONCLUSION

This study validates two efficient and robust smartphone-based methods for hearing assessment—GRaBr for pure-tone threshold estimation and rACALOS for categorical loudness scaling—under typical home conditions but with calibrated hardware. The findings demonstrate that both measures provide reliable and valid outcomes comparable to those obtained in controlled laboratory settings.

In the validation experiment, pure-tone audiometry using GRaBr and categorical loudness scaling using rACALOS yielded equivalent results between home and sound-attenuated environments at 0.25, 1, and 4 kHz, confirming the validity of remote testing. Test–retest results further indicate moderate-to-good reliability across sessions, supporting the consistency of home-based measurements.

GRaBr outperformed the SIUD method in reliability and accuracy across all tested frequencies, making it a preferred approach for mobile threshold estimation. Similarly, rACALOS produced threshold estimates that were closer to audiometric thresholds measured by SIUD and GRaBr than those from the baseline ACALOS procedure, demonstrating its advantage in improving hearing threshold estimation and test efficiency by combining threshold and loudness assessments.

Overall, the results support the feasibility of using GRaBr and rACALOS for reliable and efficient hearing assessments in real-world home environments, marking a step forward toward validated smartphone-based audiometry for large-scale or remote applications.

## Data Availability

All data produced in the present study are available upon reasonable request to the authors

## ACKNOWLEDGMENTS

This work was funded by the Deutsche Forschungsgemeinschaft (DFG, German Research Foundation) under Germany’s Excellence Strategy – EXC 2177/1 - Project ID 390895286.

## GLOSSARY

Abbreviation: Meaning
ACALOS: adaptive categorical loudness scaling
ANOVA: analysis of variance
B&K: Brüel&Kjaer
BTUX: fitting method for loudness function in ACALOS
CLS: categorical loudness scaling
CU: categorical units
FL: fixed-level procedure
GRaBr: graded response bracketing
HI: hearing impaired
HTL: hearing threshold level (at 2.5 CU on the loudness function)
ICC: intraclass cross-correlation
IQR: interquartile ranges
MEL-Med: maximum expected information-median
MEL-ML: maximum expected information-maximum likelihood
MIQR: mean interquartile range
MPANLs: maximum permissible ambient noise levels
MSD: mean signed difference
NCE: noise reduction earphones
NH: normal hearing
PTA: pure-tone average
qCLS: quick categorical loudness scaling
rACALOS: reinforced adaptive categorical loudness scaling
RMSE: root mean squared error
SA: slope-adaptive procedure
SIUD: single interval up and down
SPL: sound pressure level

## REFERENCES

1. Akeroyd, M. A., Arlinger, S., Bentler, R. A., Boothroyd, A., Dillier, N., Dreschler, W. A., … & Kollmeier, B. (2015). International Collegium of Rehabilitative Audiology (ICRA) recommendations for the construction of multilingual speech tests: ICRA Working Group on Multilingual Speech Tests. International journal of audiology, 54(sup2), 17–22.

2. Almufarrij, I., Dillon, H., Dawes, P., Moore, D. R., Yeung, W., Charalambous, A. P., … & Munro, K. J. (2022). Web-and app-based tools for remote hearing assessment: a scoping review. International Journal of Audiology, 1-14.

3. American National Standards Institute. Maximum Permissible Ambient Noise Levels for Audiometric Test Rooms. (ANSI S3.1–R2018).New York, NY: American National Standards Institute; 2018

4. Bean, B. N., Roberts, R. A., Picou, E. M., Angley, G. P., & Edwards, A. J. (2022). Automated audiometry in quiet and simulated exam room noise for listeners with normal hearing and impaired hearing. Journal of the American Academy of Audiology, 33(01), 006–013.

5. Behar, A. (2021). Audiometric tests without booths. International Journal of Environmental Research and Public Health, 18(6), 3073.

6. Bianco, R., Mills, G., de Kerangal, M., Rosen, S., & Chait, M. (2021). Reward enhances online participants’ engagement with a demanding auditory task. Trends in Hearing, 25, 23312165211025941.

7. Bland, J. M., & Altman, D. G. (1999). Measuring agreement in method comparison studies. Statistical methods in medical research, 8(2), 135–160.

8. Bland, J. M., & Altman, D. G. (2007). Agreement between methods of measurement with multiple observations per individual. Journal of biopharmaceutical statistics, 17(4), 571–582.

9. Brand, T., & Hohmann, V. (2002). An adaptive procedure for categorical loudness scaling. The Journal of the Acoustical Society of America, 112(4), 1597–1604.

10. Brand, T., 2000. Analysis and Optimization of Psychophysical Procedures in Audiology. Universität Oldenburg, Germany. PhD thesis.

11. Brennan-Jones, C. G., Eikelboom, R. H., Swanepoel, D. W., Friedland, P. L., & Atlas, M. D. (2016). Clinical validation of automated audiometry with continuous noise-monitoring in a clinically heterogeneous population outside a sound-treated environment. International journal of audiology, 55(9), 507–513.

12. Buhl, M., Akin, G., Saak, S., Eysholdt, U., Radeloff, A., Kollmeier, B., & Hildebrandt, A. (2022). Expert validation of prediction models for a clinical decision-support system in audiology. Frontiers in Neurology, 13, 960012.

13. Clark, J. G., Brady, M., Earl, B. R., Scheifele, P. M., Snyder, L., & Clark, S. D. (2017). Use of noise cancellation earphones in out-of-booth audiometric evaluations. International Journal of Audiology, 56(12), 989–996.

14. Fultz, S. E., Neely, S. T., Kopun, J. G., & Rasetshwane, D. M. (2020). Maximum expected information approach for improving efficiency of categorical loudness scaling. Frontiers in Psychology, 11, 578352.

15. Giavarina, D. (2015). Understanding bland altman analysis. Biochemia medica, 25(2), 141–151.

16. Hazan, A., Luberadzka, J., Rivilla, J., Snik, A., Albers, B., Méndez, N., … & Kinsbergen, J. (2022). Home-Based Audiometry With a Smartphone App: Reliable Results?. American Journal of Audiology, 31(3S), 914–922.

17. Hellbrück, J. (1987). How to measure loudness under natural conditions?. The Japanese Journal of Ergonomics, 23(5), 307–310.

18. Honeth, L., Bexelius, C., Eriksson, M., Sandin, S., Litton, J. E., Rosenhall, U., … & Bagger-Sjöbäck, D. (2010). An internet-based hearing test for simple audiometry in nonclinical settings: preliminary validation and proof of principle. Otology & Neurotology, 31(5), 708–714.

19. ISO 16832, 2006. Acousticsd Loudness Scaling by Means of Categories. Standard of the International Organization for Standardization, Geneva, Switzerland.

20. Kohlrausch, A., Fassel, R., Van Der Heijden, M., Kortekaas, R., Van De Par, S., Oxenham, A. J., & Püschel, D. (1997). Detection of tones in low-noise noise: Further evidence for the role of envelope fluctuations. Acta Acustica united with Acustica, 83(4), 659–669.

21. Kollmeier, B. (Ed.). (1997). Hörflächenskalierung: Grundlagen und Anwendung der kategorialen Lautheitsskalierung für Hördiagnostik und Hörgeräte-Versorgung. Median-Verlag von Killisch-Horn.

22. Kollmeier, B., Gilkey, R. H., & Sieben, U. K. (1988). Adaptive staircase techniques in psychoacoustics: A comparison of human data and a mathematical model. The Journal of the Acoustical Society of America, 83(5), 1852–1862.

23. Koo, T. K., & Li, M. Y. (2016). A guideline of selecting and reporting intraclass correlation coefficients for reliability research. Journal of chiropractic medicine, 15(2), 155–163.

24. Kopun, J. G., Turner, M., Harris, S. E., Kamerer, A. M., Neely, S. T., & Rasetshwane, D. M. (2022). Evaluation of Remote Categorical Loudness Scaling. American journal of audiology, 31(1), 45–56.

25. Kursun, Bertan & Petersen, Erik & Shen, Yi. (2023). Exploring Self-directed Hearing-aid Fitting with No Booth And No Audiogram. 10.13140/RG.2.2.19575.19360.

26. Lecluyse, W., & Meddis, R. (2009). A simple single-interval adaptive procedure for estimating thresholds in normal and impaired listeners. The Journal of the Acoustical Society of America, 126(5), 2570–2579.

27. Maclennan-Smith, F., Swanepoel, D. W., & Hall III, J. W. (2013). Validity of diagnostic pure-tone audiometry without a sound-treated environment in older adults. International journal of audiology, 52(2), 66–73.

28. Margolis, R. H., Saly, G. L., & Wilson, R. H. (2022). Ambient Noise Monitoring during Pure-Tone Audiometry. Journal of the American Academy of Audiology, 33(01), 045–056.

29. Masalski, M., Grysiński, T., & Kręcicki, T. (2014). Biological calibration for web-based hearing tests: evaluation of the methods. Journal of medical Internet research, 16(1), e2798.

30. Meinke, D. K., & Martin, W. H. (2023). Boothless audiometry: Ambient noise considerations. The Journal of the Acoustical Society of America, 153(1), 26–39.

31. Min, S. H., & Zhou, J. (2021). Smplot: An R package for easy and elegant data visualization. Frontiers in Genetics, 12, 2582.

32. Oetting, D., Brand, T., & Ewert, S. D. (2014). Optimized loudness-function estimation for categorical loudness scaling data. Hearing Research, 316, 16–27.

33. Ooster, J., Krueger, M., Bach, J. H., Wagener, K. C., Kollmeier, B., & Meyer, B. T. (2020). Speech audiometry at home: automated listening tests via smart speakers with normal-hearing and hearing-impaired listeners. Trends in Hearing, 24, 2331216520970011.

34. Peng, Z. E., Waz, S., Buss, E., Shen, Y., Richards, V., Bharadwaj, H., … & Venezia, J. H. (2022). Remote testing for psychological and physiological acoustics. The Journal of the Acoustical Society of America, 151(5), 3116–3128.

35. Rasetshwane, D. M., Trevino, A. C., Gombert, J. N., Liebig-Trehearn, L., Kopun, J. G., Jesteadt, W., … & Gorga, M. P. (2015). Categorical loudness scaling and equal-loudness contours in listeners with normal hearing and hearing loss. The Journal of the Acoustical Society of America, 137(4), 1899–1913.

36. Revelle, W. (2018). psych: Procedures for psychological, psychometric, and personality research.

37. Robler, S. K., Coco, L., & Krumm, M. (2022). Telehealth solutions for assessing auditory outcomes related to noise and ototoxic exposures in clinic and research. The Journal of the Acoustical Society of America, 152(3), 1737–1754.

38. Saliba, J., Al-Reefi, M., Carriere, J. S., Verma, N., Provencal, C., & Rappaport, J. M. (2017). Accuracy of mobile-based audiometry in the evaluation of hearing loss in quiet and noisy environments. Otolaryngology–Head and Neck Surgery, 156(4), 706–711.

39. Sanchez-Lopez, R., Nielsen, S. G., El-Haj-Ali, M., Bianchi, F., Fereczkowski, M., Cañete, O. M., … & Santurette, S. (2021). Auditory tests for characterizing hearing deficits in listeners with various hearing abilities: The BEAR test battery. Frontiers in neuroscience, 15, 724007.

40. Scharf, M. K., Huber, R., Schulte, M., & Kollmeier, B. (2024). Microphone calibration estimation for mobile audiological tests with resonating bottles. International Journal of Audiology, 1-7. DOI: 10.1080/14992027.2024.2395416

41. Serpanos, Y. C., Hobbs, M., Nunez, K., Gambino, L., & Butler, J. (2022). Adapting audiology procedures during the pandemic: Validity and efficacy of testing outside a sound booth. American Journal of Audiology, 31(1), 91–100.

42. Shen, Y., Zhang, C., & Zhang, Z. (2018). Feasibility of interleaved Bayesian adaptive procedures in estimating the equal-loudness contour. The Journal of the Acoustical Society of America, 144(4), 2363–2374.

43. Storey, K. K., Muñoz, K., Nelson, L., Larsen, J., & White, K. (2014). Ambient noise impact on accuracy of automated hearing assessment. International Journal of Audiology, 53(10), 730–736.

44. Swanepoel, D. W., Matthysen, C., Eikelboom, R. H., Clark, J. L., & Hall III, J. W. (2015). Pure-tone audiometry outside a sound booth using earphone attentuation, integrated noise monitoring, and automation. International Journal of Audiology, 54(11), 777–785.

45. Swanepoel, D. W., Mngemane, S., Molemong, S., Mkwanazi, H., & Tutshini, S. (2010). Hearing assessment—reliability, accuracy, and efficiency of automated audiometry. Telemedicine and e-Health, 16(5), 557–563.

46. Thai-Van, H., Joly, C. A., Idriss, S., Melki, J. B., Desmettre, M., Bonneuil, M., … & Reynard, P. (2023). Online digital audiometry vs. conventional audiometry: a multi-centre comparative clinical study. International Journal of Audiology, 62(4), 362–367.

47. Trevino, A. C., Jesteadt, W., & Neely, S. T. (2016). Development of a multi-category psychometric function to model categorical loudness measurements. The Journal of the Acoustical Society of America, 140(4), 2571–2583.

48. Wickham, H., Averick, M., Bryan, J., Chang, W., McGowan, L. D. A., François, R., … & Yutani, H. (2019). Welcome to the Tidyverse. Journal of open source software, 4(43), 1686.

49. Wiseman, K., Slotkin, J., Spratford, M., Haggerty, A., Heusinkvelt, M., Weintraub, S., … & McCreery, R. (2023). Validation of a tablet-based assessment of auditory sensitivity for researchers. Behavior research methods, 55(6), 2838–2852.

50. Xu, C. (2025). Crucial Elements of a Virtual Hearing Clinic on Mobile Devices - Psychophysics, Diagnostic Parameter Estimation and Validation (Doctoral dissertation, University of Oldenburg Press (UOLP)), ISBN 978-3-8142-2424-4. doi: 10.13140/RG.2.2.14023.00169

51. Xu, C., & Kollmeier, B. (2025). Calibration offset estimation in mobile hearing tests via categorical loudness scaling. arXiv preprint arXiv:2508.14824.

52. Xu, C., Hülsmeier, D., Buhl, M., & Kollmeier, B. (2024a). How Does Inattention Influence the Robustness and Efficiency of Adaptive Procedures in the Context of Psychoacoustic Assessments via Smartphone?. Trends in Hearing, 28, 23312165241288051.

53. Xu, C., Schell-Majoor, L., & Kollmeier, B. (2024b). Development and verification of non-supervised smartphone-based methods for assessing pure-tone thresholds and loudness perception. International Journal of Audiology, 1-11.

54. Zhao, S., Brown, C. A., Holt, L. L., & Dick, F. (2022). Robust and Efficient Online Auditory Psychophysics. Trends in hearing, 26, 23312165221118792.

55. Zwicker, E., & Fastl, H. (2013). Psychoacoustics: Facts and models (Vol. 22). Springer Science & Business Media.

